# Frequency of HIV serodifferent couples within TB-affected households in a setting with a high burden of HIV-associated TB

**DOI:** 10.1101/2022.11.28.22282837

**Authors:** Godwin Anguzu, Amanda J Gupta, Emmanuel Ochom, Ashley S. Tseng, Meixin Zhang, Ruanne V. Barnabas, Abraham D. Flaxman, Achilles Katamba, J. Lucian Davis, Jennifer M. Ross

## Abstract

**Background:** Strong epidemiological links between HIV and tuberculosis (TB) may make household TB contact investigation an efficient strategy for HIV screening and finding individuals in serodifferent partnerships at risk of HIV and linking them to HIV prevention services. We aimed to compare the proportions of HIV serodifferent couples in TB-affected households and in the general population of Kampala, Uganda.

**Methods:** We included data from a cross-sectional trial of HIV counselling and testing (HCT) in the context of home-based TB evaluation in Kampala, Uganda in 2016-2017. After obtaining consent, community health workers visited the homes of participants with TB to screen contacts for TB and offer HCT to household members ≥15 years. We defined index participants and their spouses and parents of the same index participant as couples, and classified couples as serodifferent if confirmed by self-reported HIV status or by HIV testing results. We used a two-sample test of proportions to compare the frequency of HIV serodifference among couples in the study to its prevalence among couples in Kampala in the 2011 Uganda AIDS Indicator Survey (UAIS).

**Results:** We included 323 index TB participants and 507 household contacts aged ≥18. Most index participants (55%) were male, while most (68%) adult contacts were female. There was ≥1 couple in 115/323 (35.6%) households, with most couples (98/115, 85.2%) including the index participant and spouse. The proportion of households with HIV-serodifferent couples was 18/323 (5.6%), giving a number-needed-to-screen of 18 households. The proportion of HIV serodifference among couples identified in the trial was significantly higher than among couples in the UAIS (15.7% vs 8%, p=0.039). The 18 serodifferent couples included 14 (77.8%) where the index participant was living with HIV and the spouse was HIV-negative, and 4 (22.2%) where the index partner was HIV-negative, while the spouse was living with HIV.

**Conclusions:** The frequency of HIV serodifference among couples identified in TB-affected households was higher than in the general population. TB household contact investigation may be an efficient strategy for identifying people with substantial exposure to HIV and linking them to HIV prevention services.

## Background

Tuberculosis (TB) is the leading cause of death in people living with human immunodeficiency virus (HIV), with an estimated 214,000 deaths resulting from TB/HIV coinfection in 2020 [1]. In Uganda, the burden of TB/HIV has been declining over time but it still remains high in 2021 with 32% of people with newly diagnosed TB also living with HIV [2–4]. TB/HIV care integration is an important element of the global End TB Strategy which seeks to reduce TB incidence and deaths by 90% and 95% respectively[5]. As the global community seeks to protect individuals in resource-constrained settings from TB/HIV[6], countries like Uganda are evaluating how contact tracing of people living with TB and/or HIV can be used to improve linkage to integrated care[7].

Evaluating household members of people with pulmonary TB for symptoms of active TB and eligibility for TB preventive treatment is a leading strategy to improve TB care and prevention. Additionally, international guidelines recommend incorporating HIV testing into TB household evaluation in settings with high HIV prevalence [8]. Ochom and colleagues demonstrated the feasibility of a community health worker (CHW)-led, home-based HIV counselling and testing approach in bridging the HIV testing gap and improving linkage to HIV care during TB household evaluation [9]. Utilizing CHWs can facilitate the integration of HIV testing and TB screening services into household settings, increasing the number of people accessing treatment and improving linkage to integrated HIV care.

A number of studies have shown that HIV-negative partners in serodifferent couples are at increased risk of acquiring HIV if the partner who is living with HIV is not taking antiretroviral therapy and virally suppressed; this observation has made HIV-seronegative partners a prioritized population for HIV pre-exposure prophylaxis (PrEP), which is a highly effective intervention to reduce the risk acquiring HIV [10–16]. Studies in Sub-Saharan Africa have shown that HIV transmission within serodifferent couples plays a major role in HIV incidence [17, 18].

Although studies have evaluated the prevalence of HIV serodifferent couples in the general population in countries in sub-Saharan Africa [19–21], there is scant literature on the frequency of HIV serodifferent couples among households in which at least one household member has been diagnosed with TB (i.e., TB-affected households). Our study evaluates the frequency of HIV serodifferent couples among TB-affected households in Kampala, Uganda and compares this to the general population to provide evidence for TB/HIV care and prevention programs.

## Methods

This analysis included data from a prospective, cross-sectional study of CHW-led HIV counselling and testing in the context of home-based TB evaluation [9]. The study was carried out in communities surrounding seven public primary care clinics providing TB services in Kampala, Uganda between July 2016 and June 2017 (Pan-African Trials Registry #20150900877140). CHWs obtained written consent from TB index participants at the clinics and visited their homes to screen contacts for TB. Additionally, participants aged 15 years or older were offered home-based HIV counseling and testing. We analyzed data from the trial to focus on the relationships described between adult (age 18 years or older) household members and the HIV status of persons within those relationships. The study was approved by the Makerere School of Medicine Research Ethics Committee, the Uganda National Council for Science and Technology, and the Yale University Human Investigation Committee.

### Relationships and defining serodifference

Relationships were classified as serodifferent if the involved persons had differences in HIV status based on HIV tests conducted during the trial or prior knowledge of being a person living with HIV. We evaluated the possible couple relationships within households using each participant’s relationship to the index participant in their household. Depending on the relationship with the index participant with TB, the contact was categorized as a spouse, parent, child, sibling, non-relative, or other relative. We did not have information about the relationship of non-index participants to other non-index members of the household.

Using the available data, we defined three types of couple relationships to the index household member with TB: 1) spouse relationship (definite couple), 2) parent relationship (probable couple), and 3) other relationship (non-informative with regard to couple status) (Table 1).

**Table 1:** Couple classification.

| **Classification** | **Title** | **Couple Description** |
| --- | --- | --- |
| **Definite** | Spouse of index participant | Index participant and a household member whose relationship to the index participant is listed as spouse. |
| **Probable** | Parents of index participant | Two household members who both list a relationship to the index participant as parent. |
| **Non-informative** | Other relationship | Unable to determine couples among participants with relationships to the index participant of child, sibling, non-relative, or other relative. |

Classification of couple relationships within households based on the relationships to the index participant with TB.

Parent relationships were characterized as probable because of the possibility to find a household with more than two parents to the TB index. Finally, other relationships were characterized as non-informative for the primary analysis because we lacked sufficient information to assign likely couples for participants whose relationship to the index participant was not as a spouse or parent. In a sensitivity analysis, we attempted to identify couples in other relationships using age groups, gender, and relationship classification, but only identified one other potential couple that was within a household that already had another couple. We did not include this couple in our primary analysis.

### Outcome definition and statistical analysis

The co-primary outcomes from this study were: 1) the frequency of HIV-serodifferent couples within TB-affected households and 2) the number of TB-affected households needed to screen to find one serodifferent couple. The first outcome was calculated as the number of households with at least one definite or probable HIV-serodifferent couple within the household divided by the number of households contacted. We also calculated the proportion of all couples identified that were serodifferent for HIV. We calculated the number of households needed to screen as the number of households screened divided by the number of households with at least one definite or probable serodifferent couple.

Participant individual and household characteristics were described using frequencies and percentages. We used a two-sample test of proportions to compare the frequency of HIV serodifference among couples in the study to versus among couples in Kampala in the 2011 Uganda AIDS Indicator Survey (UAIS), which was the most recent large, population-based survey to report HIV serodifference among couples. The UAIS was a nationally representative, population-based survey designed to obtain national and sub-national estimates of HIV prevalence, syphilis infection, and other program indicators. For the 2011 UAIS survey, a two-stage sample design was used to obtain a representative sample of 11,750 households in which interviews were conducted, including 181 couples in Kampala. Additionally, 4,724 (unweighted) cohabiting couples were both interviewed and then tested for HIV [22]. Secondly, we compared the frequency of TB-affected households with at least one member living with HIV to the prevalence of households with the same from the Uganda population-based HIV impact assessment survey (UPHIA 2016). The UPHIA was a national population-based survey that provided estimates of HIV prevalence and viral suppression at national and regional levels. In the UPHIA 2016, 12,800 households throughout Uganda were sampled, including 3,368 households in urban areas [23]. Finally, to examine bivariate associations with having a serodifferent couple within the household, we used simple proportions and Pearson Chi-square tests (P-value < 0.05).

## Results

The study included 323 index participants with TB from 323 households, with the majority (78.6%) residing in Kampala and the remainder living in nearby Wakiso district. CHWs interviewed 507 contacts of the index participants aged 18 and older. More than half (55.4%) of index participants were male, whereas most (67.5%) of the contacts were female (Table 2). The HIV frequency among index participants was 32.5% and was 12.6% among contacts ≥15 years and 0.9% among contacts <15. Seven (1.4%) of contacts were diagnosed with TB during the study. One hundred twenty-seven of the 323 TB-affected households (39.3%) included at least one member living with HIV, which was higher than the 16.6% of households from the general population in urban areas of Uganda with at least one member living with HIV in the UPHIA survey that took place during the same years (risk difference = 22.7%, 95% confidence interval (CI) = 17.2% - 28.2%, p<0.001) [23].

**Table 2.** Characteristics of index participants and household contacts ages 18 and older.

| **Characteristics** | **Index participants**<br>**323 (%)** | **Adult household contacts**<br>**507 (%)** |
| --- | --- | --- |
| **Median age years (IQR)** | 28 (22 - 36) | 25 (19 - 36) |
| **Gender** |  |  |
| **Female** | 144 (44.6) | 342 (67.5) |
| **Male** | 179 (55.4) | 165 (32.5) |
| **HIV status\*** |  |  |
| **Negative** | 216 (66.9) | 370 (73.0) |
| **Positive** | 105 (32.5) | 59 (11.6) |
| **Unknown** | 2 (0.6) | 29 (5.7) |
| **Never tested** | - | 45 (8.9) |
| **Decline to state** | - | 4 (0.8) |
| **Diagnosed with TB** |  |  |
| **Yes** | 323 (100.0) | 7 (1.4) |
| **No** | 0 | 500 (98.6) |

**Abbreviations:** IQR - Interquartile range
**Legend:** Unavailable HIV status on categories of never tested and decline to state for index participants

The most common relationship of the contact to the index participant was child (32.5%) followed by sibling (27.1%). Within 323 households, 115 (35.6%) households had at least one couple identified, with the majority of the couples 98/115 (85.2%) involving the index participant and their spouse, followed by parent relationships (**Fig. 1**). Two hundred eight (64.4%) households had no couple relationships identified.

**Figure 1.**
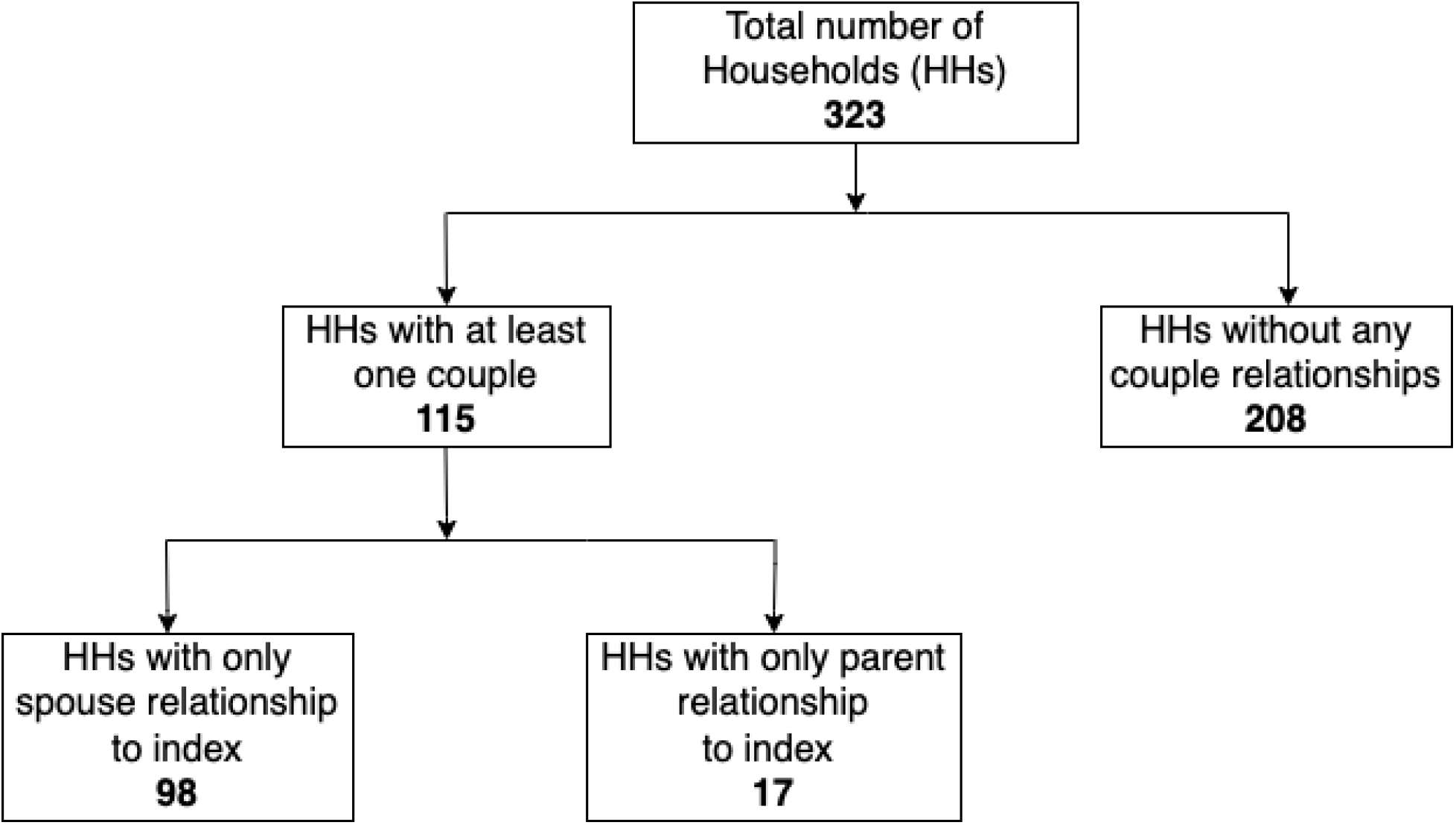
Flow diagram showing couples within households. The enrolled households included 115 households with at least one couple identified.

The proportion of TB-affected households that included at least one HIV-serodifferent couple was 18/323 (5.6%). All 18 serodifferent couples included the spouse of an index participant with TB. The number of households needed to screen to identify one HIV-serodifferent couple was 17.9. Among the 115 couples identified in TB-affected households, the frequency of HIV-serodifferent couples was 18/115 (15.7%) (Table 3).

**Table 3:**
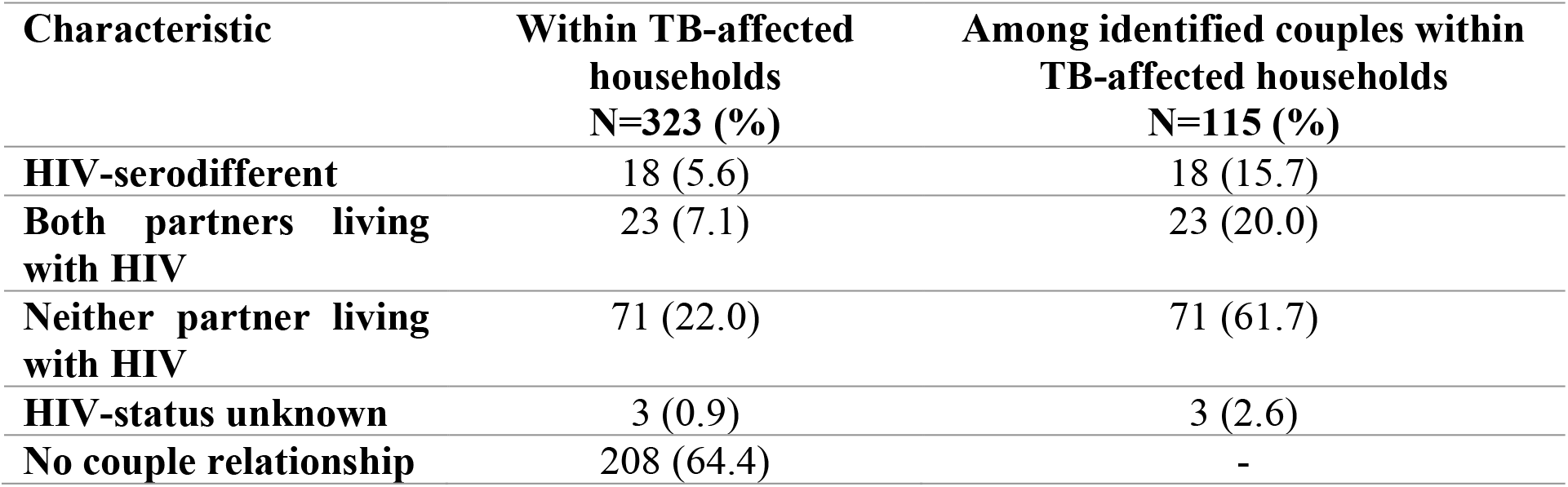
Categories of serodifference among couples living in TB affected households.

| Characteristic | Within TB-affected households<br>N=323 (%) | Among identified couples within TB-affected households<br>N=115 (%) |
| --- | --- | --- |
| **HIV-serodifferent** | 18 (5.6) | 18 (15.7) |
| **Both partners living with HIV** | 23 (7.1) | 23 (20.0) |
| **Neither partner living with HIV** | 71 (22.0) | 71 (61.7) |
| **HIV-status unknown** | 3 (0.9) | 3 (2.6) |
| **No couple relationship** | 208 (64.4) | - |

The frequency of HIV serodifference among couples identified in TB-affected households in this study was significantly higher than among couples in the general population in Kampala recorded in the Uganda AIDS Indicator Survey in 2011 (15.7% vs 8%, risk difference = 7.7%, 95% confidence interval (CI) = 0 - 15.4%, p<0.039) [21]. Among the 18 serodifferent couples, 14 (77.8%) couples were in a spouse relationship where the index partner was living with both HIV and TB while their partners were HIV-negative. Conversely, four (22.2%) couples were in a spouse relationship where the index partner with TB was HIV-negative, while their partner was living with HIV.

Table 4 shows the demographic characteristics of couples identified in TB-affected households by HIV patterns. The frequency of serodifference among women aged 18 to 29 years (15.5%) and 30 to 39 years (18.2%) was relatively similar to those aged ≥40 years (14.3%). The point estimates for frequency of serodifference were not different between men aged ≥40 years (21.4%) and 30 to 39 years (17.8%) compared to those aged 18 to 29 years (4.0%) (p-value=0.19). Men older than their partners by 5 to 9 years had a higher point estimate of frequency of serodifference (26.3%) than men who were of the same age or older by only 0 to 4 years (11.6%), though the difference was not significantly different. There was a large but non-significant serodifference in Kampala (19.8%) compared to Wakiso (6.5%) (p-value=0.058).

**Table 4:**
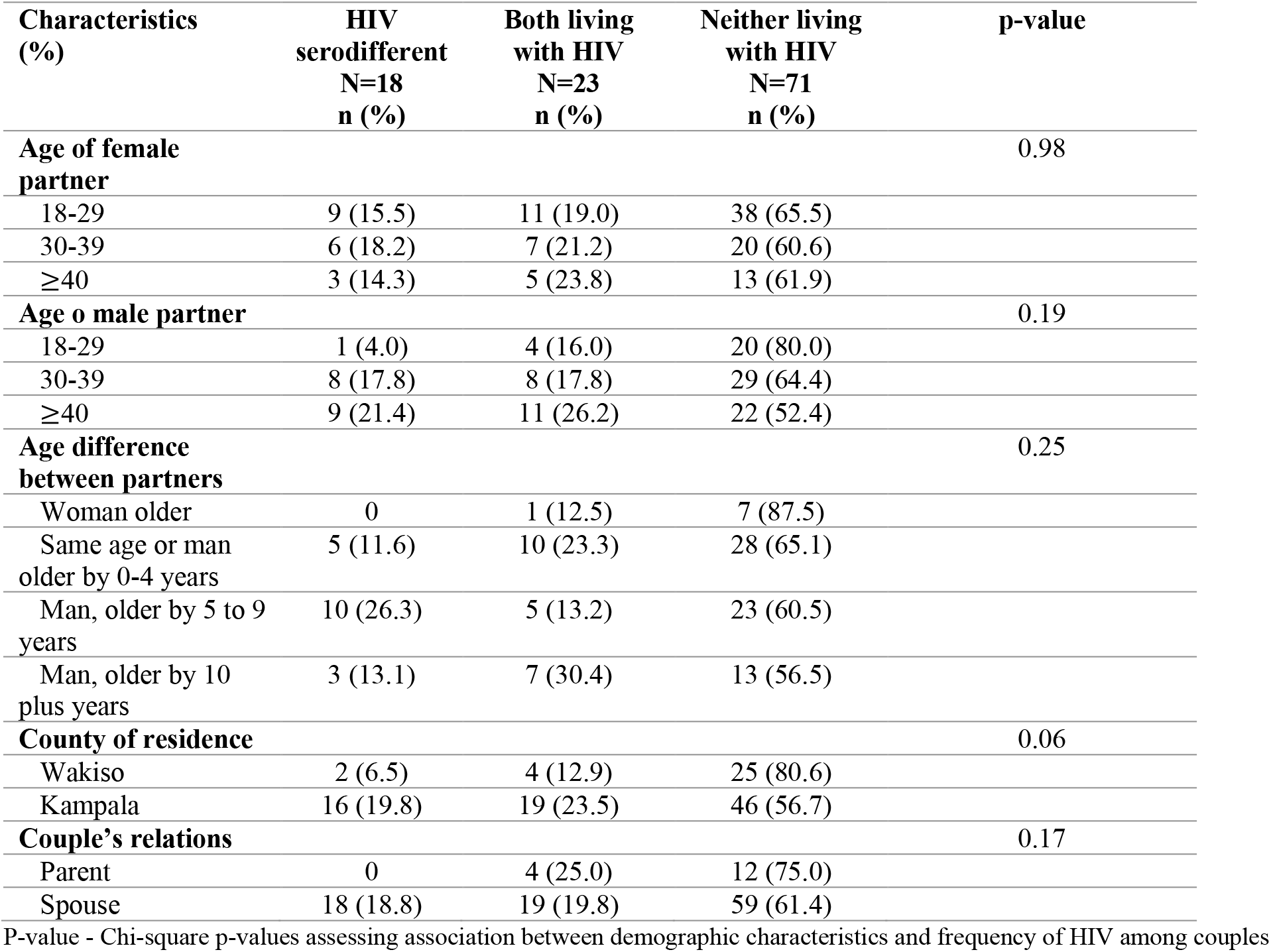
Demographic characteristics of couples living in TB-affected households by HIV patterns (N=112)

| **Characteristics (%)** | **HIV serodifferent**<br>**N=18**<br>**n (%)** | **Both living with HIV**<br>**N=23**<br>**n (%)** | **Neither living with HIV**<br>**N=71**<br>**n (%)** | **p-value** |
| --- | --- | --- | --- | --- |
| **Age of female partner** |  |  |  | 0.98 |
| 18-29 | 9 (15.5) | 11 (19.0) | 38 (65.5) |  |
| 30-39 | 6 (18.2) | 7 (21.2) | 20 (60.6) |  |
| ≥40 | 3 (14.3) | 5 (23.8) | 13 (61.9) |  |
| **Age of male partner** |  |  |  | 0.19 |
| 18-29 | 1 (4.0) | 4 (16.0) | 20 (80.0) |  |
| 30-39 | 8 (17.8) | 8 (17.8) | 29 (64.4) |  |
| ≥40 | 9 (21.4) | 11 (26.2) | 22 (52.4) |  |
| **Age difference between partners** |  |  |  | 0.25 |
| Woman older | 0 | 1 (12.5) | 7 (87.5) |  |
| Same age or man older by 0-4 years | 5 (11.6) | 10 (23.3) | 28 (65.1) |  |
| Man, older by 5 to 9 years | 10 (26.3) | 5 (13.2) | 23 (60.5) |  |
| Man, older by 10 plus years | 3 (13.1) | 7 (30.4) | 13 (56.5) |  |
| **County of residence** |  |  |  | 0.06 |
| Wakiso | 2 (6.5) | 4 (12.9) | 25 (80.6) |  |
| Kampala | 16 (19.8) | 19 (23.5) | 46 (56.7) |  |
| **Couple's relations** |  |  |  | 0.17 |
| Parent | 0 | 4 (25.0) | 12 (75.0) |  |
| Spouse | 18 (18.8) | 19 (19.8) | 59 (61.4) |  |

P-value - Chi-square p-values assessing association between demographic characteristics and frequency of HIV among couples

## Discussion

We calculated the frequency of HIV serodifferent couples among participants in a trial of TB household contact evaluation that included household-based HIV testing in Uganda to inform a potential strategy for integrating HIV prevention with TB household contact investigation. We identified couple relationships in just over one-third of households, among whom the proportion with HIV-serodifference was nearly two times higher than among couples from the general population of Kampala in the 2011 AIS (15.7% vs 8.0%). However, with the relatively low proportion of households in which a couple was identified, the proportion of TB-affected households in which an HIV-serodifferent couple was identified was also low at 5.6%. We did not find any significant demographic differences between serodifferent and seroconcordant couples, although we may not have had sufficient power to characterize these differences.

PrEP is highly effective for preventing HIV acquisition among people without HIV whose partners are living with HIV and are not virally suppressed, and strategies are needed to increase PrEP uptake among populations who would benefit [24]. Our study supports a potential benefit for integrating HIV prevention with TB household evaluation because the frequency of HIV serodifference among couples identified in our study was higher than among couples in the general population of Kampala from the UAIS. Comparing to other settings, the prevalence of HIV serodifference among couples estimated from a population-based survey in Kenya was 5.8% [25]. In addition, four prevalence surveys conducted in Ndhiwa (Kenya), Chiradzulu (Malawi), Gutu (Zimbabwe) and Nsanje (Malawi), found an overall prevalence of 10.9% [26]. These estimates were comparable to the population-based AIS study in Uganda and lower than the estimate from our study. The high estimate in the current study in relation to the population-based studies may suggest the relevance of home-based TB contact tracing to identify couples engaging in HIV-serodifferent relationships.

While HIV testing studies often focus on the yield of people newly found to be living with HIV, our study expands the framework for valuing home-based testing by identifying people who may have substantial HIV exposure and may benefit from PrEP. More than one-third of households in our study included at least one household member living with HIV, which indicates an opportunity to offer linkage to ART and/or support for ART adherence during TB household contact evaluation. Assessing HIV exposures beyond serodifferent partnerships within the household may also increase the value of testing.

Unique to this study, the proportion of TB-affected households with an HIV serodifferent couple was 5.6%. Within the 18 serodifferent couples, 14 couples included one partner living with both active TB and HIV (with partners who did not have TB or HIV), and 4 couples included one partner living with active TB without HIV and the other partner living with HIV without active TB. These two patterns highlight the potential for transmission of either HIV or TB within couples and the importance of screening and linkage to integrated TB/HIV care.

We did not find any significant demographic differences between serodifferent and seroconcordant couples. We had limited power to detect differences due to the small number of serodifferent couples in the study. In UAIS, serodifference was more common among couples where the male partner was at least 10 years older than the female partner [22]. Additionally, we found a lower proportion of couples where both members were HIV-negative in our study (62.1%) compared to 90.3% in UAIS. In other settings, age 35-45 years for women and men, rare condom use and active TB at study baseline were associated with serodifference among couples. On the other hand, women with older male partners and couples living far away from community health centers versus those living closure were less likely to be engaged in HIV-serodifferent relationships [27].

Our study had several strengths. To our knowledge, this is among the first studies published that characterizes HIV-serodifferent couples among TB-affected households in a high prevalence setting. Even though there might have been some random misclassification of participants not directly estimated, only three couples in our study had unknown HIV status. Our study reduces the knowledge gap around a potential missed opportunity for HIV prevention in the setting of TB household contact investigation, which is a is a pillar of the End TB Strategy and the UNAIDS Strategy 2016-2021 [5].

Our study had limitations. First, we identified a small number of couples within households (n=115), limiting the power of our study and ability to detect significant differences by demographic characteristics of serodifferent couples. We likely undercounted the number of couples in these households because the available data from the study recorded the relationship of each person in the household to the index participant with TB, but not the relationship of every person to each other. This limitation could bias our estimate of the frequency of serodifferent couples within households downward and could bias upward our estimate of the proportion of couples who were serodifferent because the index participants with TB were more likely to be living with HIV than other members of the household. Second, we did not have data to characterize couples where one partner lived outside the household. Third, we had limited data on ART use for individuals living with HIV, which impacts the potential benefit of PrEP to prevent HIV transmission.

## Conclusions

Among couples identified in households affected by TB, the frequency of HIV serodifference was higher than among couples in the general population of urban residents, though we had limited ability to identify couple relationships that did not include the index participant with TB. TB household contact investigation may be an efficient strategy for identifying people with substantial exposure to HIV and linking them to HIV prevention services.

## Data Availability

The datasets used and/or analyzed during the current study are available from the corresponding author on reasonable request.

## Declarations

### Ethics approvals and consent to participate

The study was conducted in accordance with the Declaration of Helsinki. The study was approved by the Makerere School of Public Health Research Ethics Committee (SPH-2021-120) and determined to be exempt research by the Yale University Human Investigation Committee (Protocol ID 2000030756).

### Consent for publication

Not applicable

### Authors’ contributions

G.A., A.K., R.V.B., J.L.D., and J.M.R designed the research study. A.J.G. and E.O. collected and curated the data. G.A. analysed the data. G.A. wrote the first draft of the manuscript and all authors contributed to subsequent versions. All authors have read and approved the final manuscript.

## Acknowledgements

The authors thank the study participants and community health workers who aided in the study.

